# Occupation, Work-Related Contact, and SARS-CoV-2 Anti-Nucleocapsid Serological Status: Findings from the Virus Watch prospective cohort study

**DOI:** 10.1101/2021.05.13.21257161

**Authors:** Sarah Beale, Parth Patel, Alison Rodger, Isobel Braithwaite, Thomas Byrne, Wing Lam Erica Fong, Ellen Fragaszy, Cyril Geismar, Jana Kovar, Annalan MD Navaratnam, Vincent Nguyen, Madhumita Shrotri, Robert W Aldridge, Andrew C Hayward, on behalf of Virus Watch Collaborative

## Abstract

**Background:** Workers differ in their risk of acquiring SARS-CoV-2 infection according to their occupation; however, few studies have been able to control for multiple confounders or investigate the work-related factors that drive differences in occupational risk. Using data from the Virus Watch community cohort study in England and Wales, we set out to estimate the total effect of occupation on SARS-CoV-2 serological status, whether this is mediated by frequency of close contact within the workplace, and how exposure to poorly ventilated workplaces varied across occupations.

**Methods:** We used data from a sub-cohort (n =3761) of adults (≥18) tested for SARS-CoV-2 anti-nucleocapsid antibodies between 01 February-28 April 2021 and responded to a questionnaire about work during the pandemic. Anti-nucleocapsid antibodies were used as a proxy of prior natural infection with COVID-19. We used logistic decomposition to estimate the total and direct effect of occupation and indirect effect of workplace contact frequency on odds of seropositivity, adjusting for age, sex, household income and region. We investigated the relationship between occupation and exposure to poorly-ventilated workplace environments using ordinal logistic regression.

**Results:** Seropositivity was 16.0% (113/707) amongst workers with daily close contact, compared to 12.9% (120/933) for those with intermediate-frequency contact and 9.6% (203/2121) for those with no work-related close contact. Healthcare (OR= 2.14, 95% CI 1.47,3.12), indoor trade, process and plant (2.09, 1.31,3.33), leisure and personal service (1.96, 1.004,3.84), and transport and mobile machine (2.17, 1.12,4.18) workers had elevated total odds of SARS-CoV-2 seropositivity compared to other professional and associate occupations. Frequency of workplace contact accounted for a variable part of the increased odds in different occupational groups (OR range 1.04 [1.0004,1.07] - 1.22 [1.07, 1.38]). Healthcare workers and indoor trades and process plant workers continued to have raised odds of infection after accounting for work-related contact, and also had had greater odds of frequent exposure to poorly-ventilated workplaces (respectively 2.15 [1.66, 2.79] and (1.51, [1.12, 2.04]).

**Discussion:** Marked variations in occupational odds of seropositivity remain after accounting for age, sex, region, and household income. Close contact in the workplace appears to contribute substantially to this variation. Reducing frequency of workplace contact is a critical part of COVID-19 control measures.

## Introduction

Occupation is a major determinant of health ^1^. This relationship has played out in real-time during the COVID-19 pandemic with people’s ability to work from home, practice social distancing, limit contact with potentially infectious individuals and work in well-ventilated conditions all shaped by their occupation ^2,3,4^.

Substantial occupational differences in severe illness and mortality have emerged in the UK and worldwide. UK Biobank data suggested possible higher relative risk of severe COVID-19 for health and social care workers during the first pandemic wave compared to non-essential workers when adjusted for age, sex, ethnicity, and country of birth^5^; however, occupational status was ascertained at study baseline (2006-2010) and may not fully account for changes in employment status. Official mortality data in the UK and Massachusetts USA suggest higher age-standardised mortality rates for workers in caring, personal service, food preparation and service, process and plant, and transportation occupations compared to the general population ^6–8^ or workers overall ^3,9^. Transport workers – specifically bus and taxi drivers - also demonstrated elevated risk of death during the first pandemic wave in Sweden compared to other professions^10^. Healthcare support workers, social care workers, construction and maintenance workers, and protective service workers also demonstrated excess mortality in the USA data^3,9^. These occupations are patient- or public-facing and/or require work outside the home^2,3,4^, potentially increasing SARS-CoV-2 exposure risk. Occupational differences in severe illness and COVID-19 mortality may be related to workplace exposure, but this cannot be easily inferred from such studies.

Emerging evidence suggests that infection risk varies substantially across occupations. Rates of antigen test positivity and nucleocapsid antibody seropositivity in healthcare workers - who are at high risk of contact with infectious individuals ^3^ and had greater access to SARS-CoV-2 testing than the general population early in the pandemic ^2^ - usually exceed those observed in the general population but vary across study populations ^11–18^. However, these estimates were largely drawn from the initial phase of the pandemic, when access to personal protective equipment (PPE) was most variable. Contact tracing during early COVID-19 outbreaks across Hong Kong, Japan, Singapore, Taiwan, Thailand, and Vietnam ^19^ suggests workplaces were a common plausible location of transmission and that occupational groups with the highest proportions of cases included healthcare, transport, sales and service, cleaning and domestic, and public safety workers. National routine testing in the Netherlands found higher antigen test positivity between June-October 2020 in hospitality workers, transport workers, and hairdressers and aestheticians compared to ‘non-close-contact’ occupations ^20^. Lower test positivity found in education and healthcare professionals compared to ‘non-close-contact’ occupations was attributed to higher testing rates in these groups as well as precautionary measures and/or PPE in these settings ^20^.

Studies investigating occupational differences in COVID-19 risk based on only the first pandemic wave, which comprises most of the existing literature, should be interpreted with caution due to changes in differential risk by occupation across pandemic waves. In the UK, employer-submitted reports of COVID-19 cases plausibly linked to the workplace ^21,22^ primarily involved health and social care workers in the first pandemic wave compared with education and manufacturing workers in the second pandemic wave. Although reporting bias may have influenced these findings, patient-facing healthcare and care workers participating in a randomly-sampled antigen testing study demonstrated higher rates of current infection than other workers in May 2020 but at no subsequent round of monthly testing up to November 2020 ^13^. A serosurvey of the entire Norwegian population over 20 years of age ^23^ similarly found greater odds of seropositivity in the first wave for healthcare and transport workers compared to the overall adult population, and in the second wave for food service workers, transport workers, and travel stewards, adjusting for age, sex, testing behaviour, and maternal country of birth. These temporal changes likely reflect the impact of widespread community transmission and of changing employment and ‘lockdown’ measures.

Few current estimates of differential infection risk across occupations are adjusted for potential socio-demographic confounding, such as the effect of deprivation. Furthermore, epidemiological investigation into the mechanisms underlying occupational differences in SARS-CoV-2 infection risk, required to inform evidence-based public health interventions, is lacking. The UK ONS Coronavirus Infection survey found little evidence of differential risk of antigen test positivity across occupations (01 Sep 2020-7 Jan 2021) after adjusting for a range of sociodemographic factors and the ability to work from home and socially distance at work ^24^. While the study tentatively concluded that contact driven by workplace attendance and ability to socially distance is likely an important driver of occupational differences, this hypothesis could not be directly tested and disaggregated from the effects of other sociodemographic factors.

Building on this indirect evidence, we aimed to address gaps in the literature around occupational differences in SARS-CoV-2 infection risk using data from the Virus Watch ^25^ community cohort study based across England and Wales. Specific research questions were:

1. How do odds of SARS-CoV-2 anti-nucleocapsid seropositivity vary across occupations? (primary objective)
2. Does frequency of work-related close contact mediate the relationship between occupation and seropositivity? (primary objective)
3. How does exposure to poorly ventilated environments vary across occupations? (secondary objective)

## Methods

### Participants

Participants in the current study were a subset of the Virus Watch study cohort. Virus Watch is a community prospective cohort study of acute respiratory infection syndromes and SARS-CoV-2 infection in England and Wales (*n*=50416 as of 28 April 2021). The study includes weekly reporting of symptoms, testing and vaccination status, as well as detailed monthly questionnaires around sociodemographic, health-related, and psychosocial/behavioural factors. The eligibility criteria, recruitment strategy, aims, and procedures for the Virus Watch study have been described in detail elsewhere ^25^, with relevant elements for the present study outlined here.

Participants in the present study comprised adults (≥18 years) who conducted monthly at-home antibody testing using self-administered capillary blood sampling kits sent for laboratory testing via post in addition to completing the main study surveys. Participants were eligible for inclusion in the present study if:

1. they self-reported their occupation upon study registration,
2. they had a valid antibody test result conducted between 01 Feb 2021 and 28 April 2021,
3. they responded to the February 2021 monthly survey regarding features of work during the pandemic.

### Exposure

Occupation, the exposure of interest, was derived from free-text job titles and descriptions in the Virus Watch baseline registration survey. Responses were semi-automatically coded using Cascot Computer Aided Structured Coding Tool Desktop Version 5.6.3^26^, the UK Office of National Statistics (ONS) recommended software for coding free-text occupational data ^27^ to generate UK Standard Occupational Classification 2020 (SOC-2020) codes ^28^.

SOC-2020 codes are grouped within minor, sub-major, and major categories based on occupation-related skill level and skill specialisation^29^. As these groupings combine occupations that take place in varying environments and with different work-related behaviour/public exposure patterns, we categorised SOC-2020 codes into the following occupational categories to reflect these factors while retaining, where possible, the occupational groups outlined by ONS: administrative and secretarial occupations; healthcare occupations; indoor trade, process & plant occupations; leisure and personal service occupations; managers, directors, and senior officials; outdoor trade occupations; sales and customer service occupations; social care and community protective services; teaching education and childcare occupations; transport and mobile machine operatives; and other professional and associate occupations (professional and associate professional occupations excluding healthcare, teaching, and social care/community protective services).

Please see Supplementary Table 1 for UK 2020 SOC codes included within each occupational category, and the most prevalent SOC-2020-defined occupations within each category.

### Outcome

The primary outcome of interest was binary-coded serological status (positive versus negative) for SARS-CoV-2 anti-nucleocapsid antibodies acquired through natural infection. Serological status was determined based on self-administered capillary samples which were sent via post for laboratory testing using the Roche Elecsys anti-N total immunoglobulin assay for the Nucleocapsid protein. Serological status was defined based on a cut-off index of ≥0.1 indicating seropositivity. Participants who provided samples across multiple months were coded as seronegative if all samples were below the cut-off value or as seropositive if any sample was above the cut-off value.

The secondary outcome of interest was frequency of workplace exposure to poor ventilated environments. This outcome was based on participants’ response to the following question in the February monthly survey: “How often does [name][surname] work indoors in an environment that is never or rarely ventilated (windows or doors opened to let in fresh air or mechanical ventilation system)?”. Responses were classified as Never (never/not applicable), Intermediate (once a month or more - once a week or more but not every day) and Every Day.

### Potential Mediator

Frequency of work-related close contact with other individuals was investigated as a potential mediator of the relationship between occupation and serological status. Contact frequency was classified as follows according to participants’ response to the following question in the 17 February monthly survey: “How often does [name][surname]’s work require close contact with others (within 2 meters, including with precautions)?”: Never (never/not applicable e.g. work from home), Intermediate (once a month or more - once a week or more but not every day) and Every Day. This question was displayed to participants who reported being in full- or part-time employment or self-employment at the time of the survey.

### Covariates

We identified potential confounders based on a purpose-developed directed acyclic graph of the relationship between occupation, work-related contact, and SARS-CoV-2 infection risk (Supplementary Figure 1) as well as the VanderWeele principle of confounder selection^30^. The following covariates were included to provide a minimally-adjusted unbiased estimate of the total and direct effects of occupation, with data drawn from the Virus Watch baseline registration questionnaire: age (<25, 30-39, 40-49, 50-59, 60+ years), sex at birth, geographic region (ONS national region), and deprivation based on annual household income (£0-24,900, £25,000-£49,999, £50,000-£75,000, and £75,000+). Based on our directed acyclic graph, the effects of other key socio-demographic confounders - such as ethnicity and underlying health conditions - were addressed through the covariates included in our analyses (see Supplementary Figure 1).

### Statistical Analyses

For all analyses, the ‘Other professional and associate’ category - which was the most prevalent occupational group in the sample (Table 1) and the group with the highest frequency of never reporting close contact in the workplace (Supplementary Table 2) - was used as the reference category (see Supplementary Table 1 for most prevalent occupations within this group). We investigated the total effect of occupation on serological status and potential mediation by frequency of work-related close contact controlling for age, sex, region and household income (see Figure 1), we applied the Buis ^31,32^ logistic decomposition method using the *ldecomp* command in Stata Version 16. Unlike standard mediation procedures, this method is able to decompose the total effect of a categorical exposure on a binary outcome with an ordinal categorical mediator, and express the total, direct and indirect effects in terms of odds ratios.

**Table 1.** Demographic Features of Participants.

| Characteristic | N = 3,761<sup>1</sup> |
| --- | --- |
| **Age** |  |
| <30 | 206 (5.5%) |
| 30-39 | 453 (12%) |
| 40-49 | 761 (20%) |
| 50-59 | 1,283 (34%) |
| 60+ | 1,058 (28%) |
| **Sex** |  |
| Female | 2,112 (56%) |
| Male | 1,642 (44%) |
| Unknown | 7 (0.2%) |
| **Occupation** |  |
| Administrative & Secretarial | 496 (13%) |
| Healthcare | 319 (8.5%) |
| Indoor Trades, Process & Plant | 247 (6.6%) |
| Leisure & Personal Service | 144 (3.8%) |
| Managers, Directors & Senior Officials | 273 (7.3%) |
| Other Professional & Associate | 1313 (35.0%) |
| Outdoor Trades | 90 (2.4%) |
| Sales & Customer Service | 175 (4.7%) |
| Social Care & Community Protective Services | 180 (4.8%) |
| Teaching, Education & Childcare | 446 (12%) |
| Transport & Mobile Machine | 78 (2.1%) |
| **Ethnicity** |  |
| White British | 3,286 (87%) |
| White Irish | 41 (1.1%) |
| White Other | 260 (6.9%) |
| Black | 16 (0.4%) |
| Mixed | 47 (1.2%) |
| Other Asian | 25 (0.7%) |
| Other Ethnicity | 13 (0.3%) |
| South Asian | 64 (1.7%) |
| Unknown | 9 (0.2%) |
| **Household Income** |  |
| £0-£24,999 | 530 (14%) |
| £25,000-£49,999 | 1,166 (31%) |
| £50,000-£74,999 | 897 (24%) |
| £75,000+ | 948 (25%) |
| Unknown | 220 (5.8%) |
| **Region** |  |
| East Midlands | 321 (8.5%) |
| East of England | 865 (23%) |
| London | 549 (15%) |
| North East | 150 (4.0%) |
| North West | 399 (11%) |
| South East | 793 (21%) |
| South West | 239 (6.4%) |
| Wales | 70 (1.9%) |
| West Midlands | 187 (5.0%) |
| Yorkshire and The Humber | 188 (5.0%) |

<sup>1</sup>n (%)

**Figure 1.**
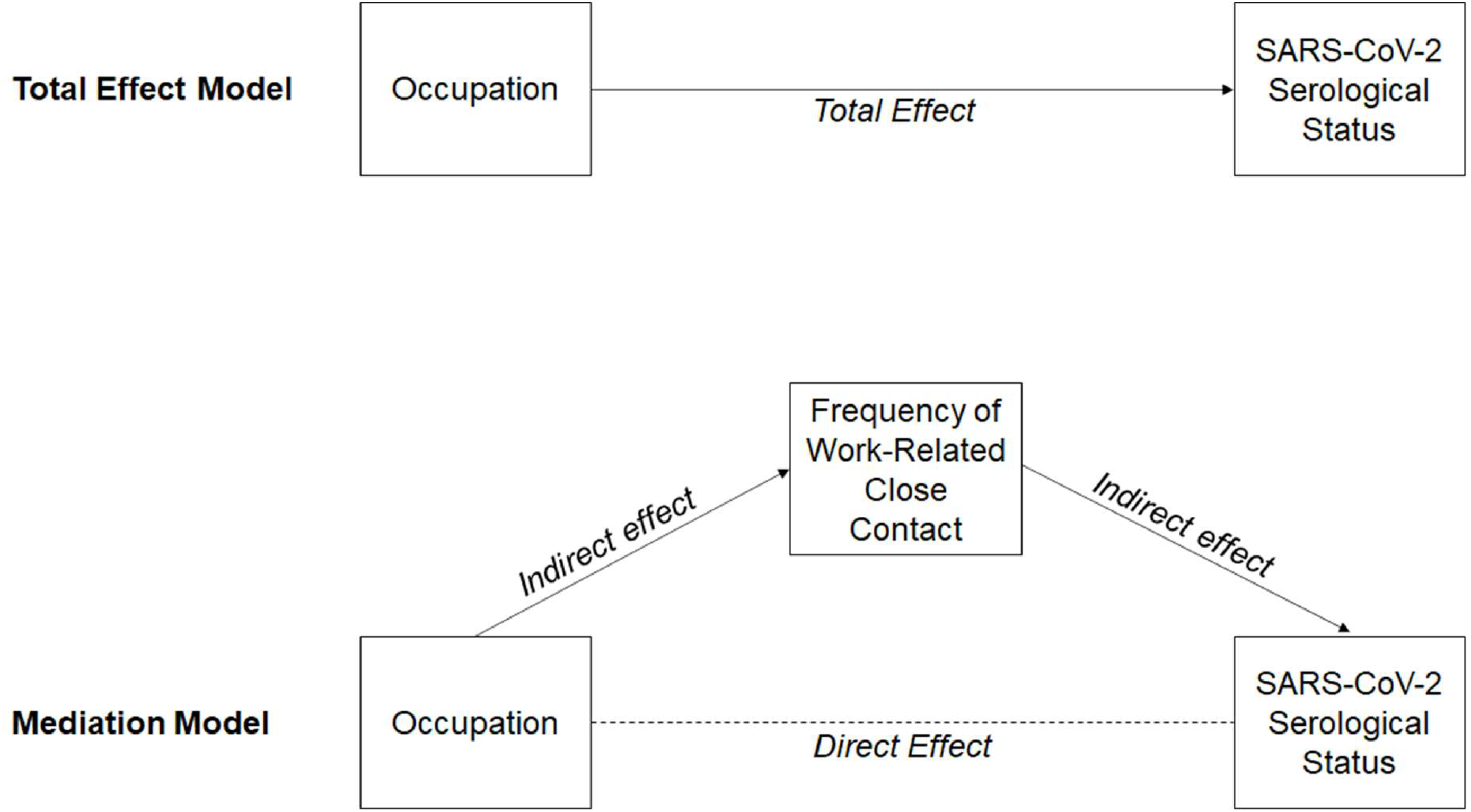
Total Effect and Mediation Models.

We applied ordered logistic regression to investigate the relationship between occupation and frequency of exposure to poorly ventilated workplaces. While poor workplace ventilation is a plausible moderator of the indirect effect of workplace contact (i.e. frequent close contact in poorly ventilated workplace environments making infection more likely), it was not included in a moderated-mediation model as it was not possible to determine if the close contacts reported also occurred in poorly ventilated spaces. This model was not adjusted for sociodemographic factors as a relationship between these factors and exposure to poorly ventilated workspaces was assumed to occur due to occupation.

There were no missing data for occupation, workplace contact frequency, age, or national region. Minimal data were missing for workplace exposure to poor ventilation (0.7%, *n*=27), sex (0.2%, *n*=7), and ethnicity (0.2%, *n*=9). Household income was missing for 5.8% of participants (*n*=220); available data were entered into models.

### Ethics and Consent

The Virus Watch study was approved by the Hampstead NHS Health Research Authority Ethics Committee: 20/HRA/2320, and conformed to the ethical standards set out in the Declaration of Helsinki. All participants provided informed consent for all aspects of the study.

## Results

Selection of participants for inclusion in the present study is illustrated in Figure 2. Table 1 reports demographic features of Virus Watch cohort participants included in the present study (*n*=3761).

**Figure 2.**
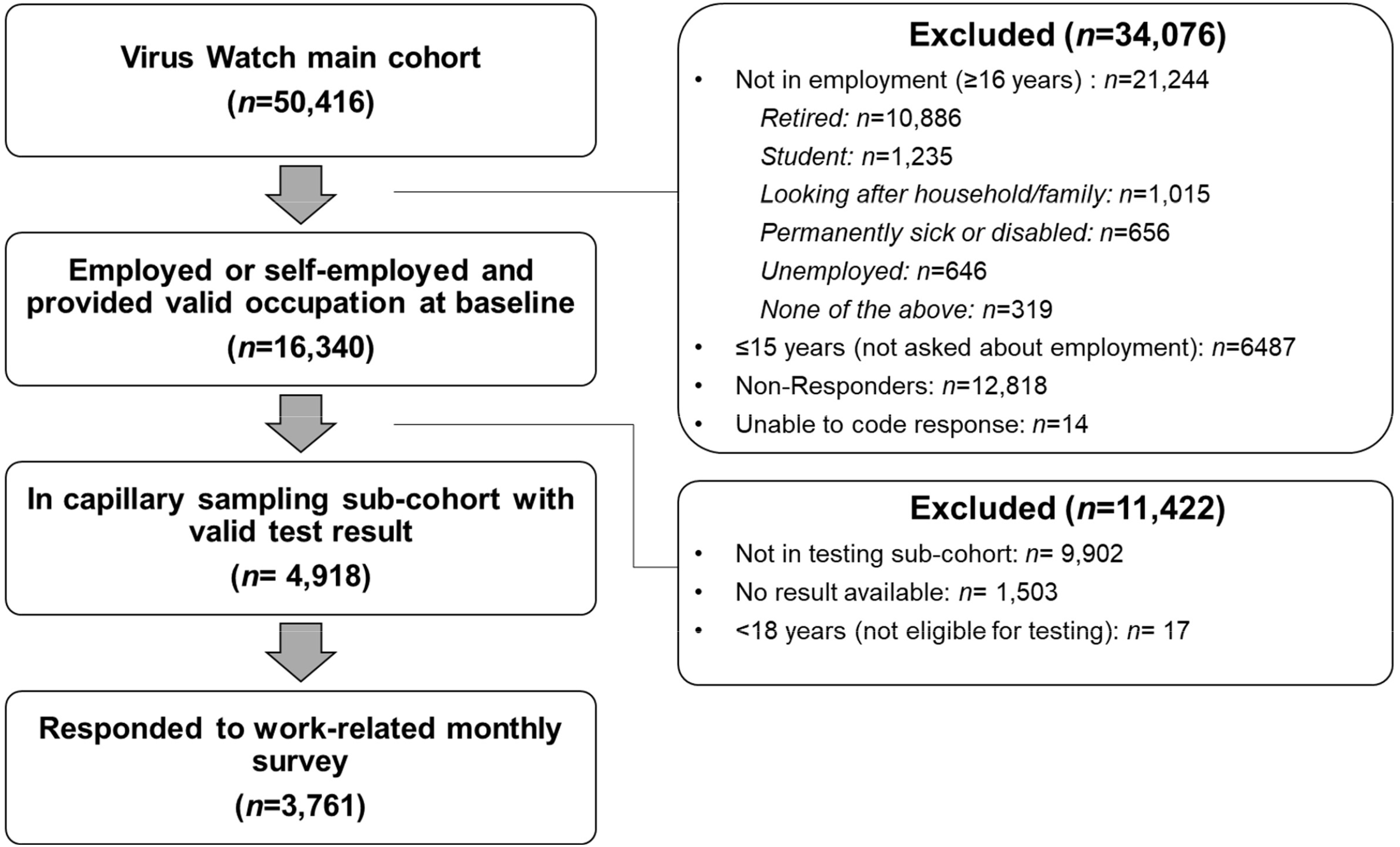
Flow Diagram of Participant Eligibility.

### Total Effect of Occupation on SARS-CoV-2 Serological Status

The proportion of seropositive and seronegative participants by occupation is reported is Table 2. Logistic regression (Figure 3 and Table 3 [total effect]) found that participants employed in healthcare professions (OR= 2.14, 95% CI 1.47,3.12), indoor trade, process and plant occupations (OR=2.09, 95%CI 1.31,3.33), leisure and personal service occupations (OR=1.96, 95% CI 1.004,3.84), and transport and mobile machine operatives (OR=2.17, 95% CI 1.12,4.18) had greater total odds of SARS-CoV-2 seropositivity compared to participants in the ‘Other professional and associate’ category.

**Table 2.** SARS-CoV-2 Serological Status by Occupation.

| Characteristic | No, N = 3,325<sup>1</sup> | Yes, N = 436<sup>1</sup> |
| --- | --- | --- |
| Occupation |  |  |
| Administrative & Secretarial | 441 (88.9%) | 55 (11.1%) |
| Healthcare | 261 (81.8%) | 58 (18.2%) |
| Indoor Trades, Process & Plant | 210 (85.0%) | 37 (15.0%) |
| Leisure & Personal Service | 121 (84.0%) | 23 (16.0%) |
| Managers, Directors & Senior Officials | 244 (89.4%) | 29 (11.6%) |
| Other Professional & Associate | 1,190 (90.6%) | 123 (9.4%) |
| Outdoor Trades | 76 (84.4%) | 14 (15.6%) |
| Sales & Customer Service | 155 (89.6%) | 20 (11.4%) |
| Social Care & Community Protective Services | 162 (90.0%) | 18 (10.0%) |
| Teaching, Education & Childcare | 399 (89.5%) | 47 (10.5%) |
| Transport & Mobile Machine | 66 (84.6%) | 12 (15.4%) |

<sup>1</sup>n (row %)

**Figure 3.**
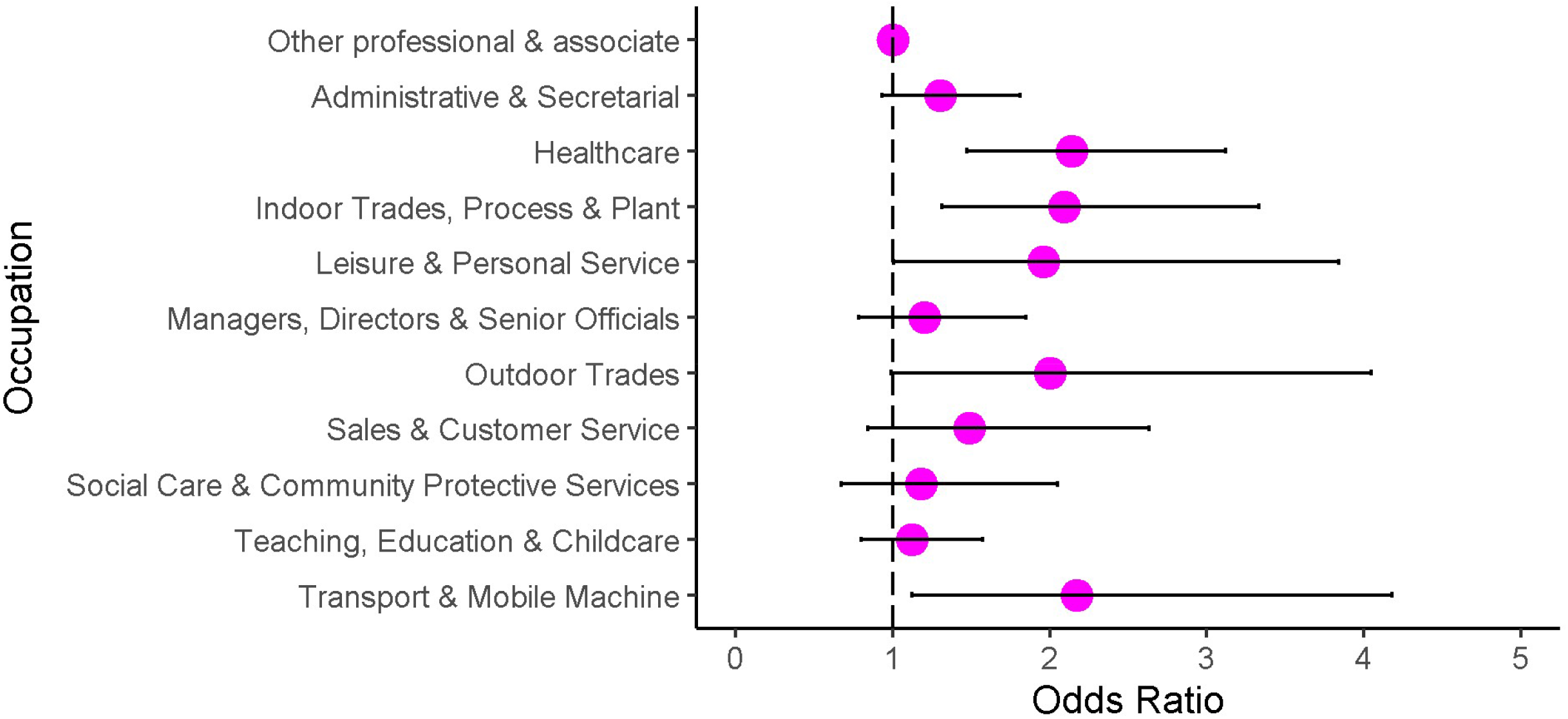
Odds of SARS-CoV-2 Seropositivity by Occupation (Total Effect Adjusted for Age, Sex, Region, and Household Income)

**Table 3.** Odds Ratios for Total, Indirect, and Direct Effects.

|  | Total |  |  | Indirect |  |  | Direct |  |  |
| --- | --- | --- | --- | --- | --- | --- | --- | --- | --- |
|  | OR | 95% CI | *p* | OR | 95% CI | *p* | OR | 95% CI | *p* |
| Other Professional & Associate | REF | REF | REF | REF | REF | REF | REF | REF | REF |
| Administrative & Secretarial | 1.3 | 0.93,1.81 | 0.12 | 1.05 | 1.01,1.09 | 0.02 | 1.24 | 0.89,1.73 | 0.20 |
| Healthcare | 2.14 | 1.47,3.12 | <0.001 | 1.21 | 1.07,1.37 | 0.002 | 1.76 | 1.20,2.59 | 0.004 |
| Indoor Trades, Process & Plant | 2.09 | 1.31,3.33 | 0.002 | 1.16 | 1.06,1.27 | 0.002 | 1.80 | 1.14,2.85 | 0.01 |
| Leisure & Personal Service | 1.96 | 1.004,3.84 | 0.049 | 1.13 | 1.05,1.23 | 0.003 | 1.73 | 0.89,3.35 | 0.10 |
| Managers, Directors & Senior Officials | 1.20 | 0.78,1.85 | 0.42 | 1.04 | 1.004,1.07 | 0.03 | 1.15 | 0.75,1.77 | 0.52 |
| Outdoor Trades | 2.00 | 0.99,4.05 | 0.05 | 1.13 | 1.04,1.23 | 0.003 | 1.76 | 0.85,3.65 | 0.13 |
| Sales & Customer Service | 1.49 | 0.84,2.63 | 0.17 | 1.10 | 1.03,1.18 | 0.004 | 1.35 | 0.74,2.44 | 0.33 |
| Social Care & Community Protective Services | 1.18 | 0.67,2.05 | 0.57 | 1.12 | 1.04,1.20 | 0.003 | 1.05 | 0.60,1.85 | 0.86 |
| Teaching, Education & Childcare | 1.12 | 0.80,1.57 | 0.52 | 1.11 | 1.03,1.20 | 0.005 | 1.00 | 0.72,1.40 | 0.98 |
| Transport & Mobile Machine | 2.17 | 1.12,4.18 | 0.02 | 1.22 | 1.07,1.38 | 0.002 | 1.78 | 0.94,3.37 | 0.08 |

#### Mediation Analysis for Workplace Contact Frequency

Workplace contact frequency by occupation is reported in Supplementary Table 2. Anti-nucleocapsid seropositivity was 16.0% (113/707) among those who had daily close contact with others at work, compared to 12.9% (120/933) among those with intermediate frequency contact and 9.6% (203/2121) among those who worked from home or never had close contact with others at work (Supplementary Table 2). Results of the models for the indirect and the direct effects are reported in Table 3. There were positive indirect effects (i.e. OR>1.00) with bootstrapped confidence intervals that did not include the value one across occupational groups, suggesting mediation of the occupation-seropositivity relationship by work-related contact frequency (OR range 1.04 [95% CI 1.0004,1.07] - 1.22 [95% CI 1.07, 1.38]). After accounting for the indirect effect of workplace contact frequency, a positive direct effect of occupation on serological status remained for healthcare professions (OR 1.76, 95% CI 1.20, 2.59), and indoor trade, process and plant occupations (OR 1.80, 95% CI 1.14, 2.85) (Table 3).

#### Exposure to Poorly Ventilated Environments by Occupation

Frequency of exposure to poorly ventilated environments by occupation is illustrated in Figure 4, with corresponding proportions in Supplementary Table 3. Anti-nucleocapsid seropositivity was 16.7% (67/401) among those who had daily exposure to poorly ventilated workplaces, compared to 12.9% (62/497) of those with intermediate exposure and 9.6% (303/2836) of those who never had exposure to poorly ventilated workplaces (Supplementary Table 3). Ordered logistic regression (Figure 5 and Supplementary Table 4) indicated that - compared to participants in the ‘Other professional and associate’ category - participants employed in healthcare professions (OR= 2.15, 95% CI 1.66, 2.79), indoor trade, process and plant occupations (OR =1.51, 95% CI 1.12, 2.04) and sales and customer service occupations (OR = 1.54, 95% CI 1.09, 2.18) had elevated odds of more frequent exposure to poorly ventilated workplace environments.

**Figure 4.**
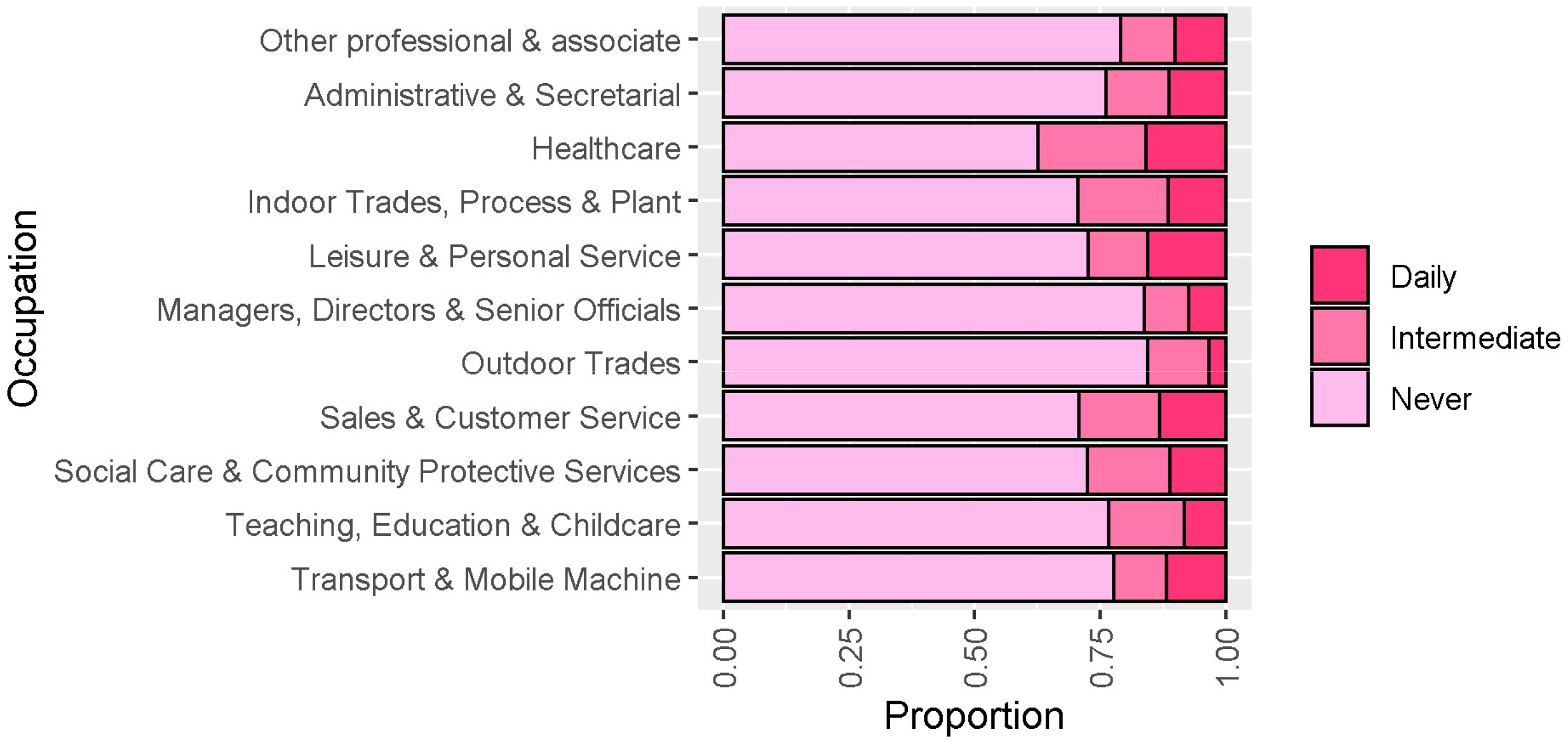
Frequency of Exposure to Poorly Ventilated Workplace by Occupation.

**Figure 5.**
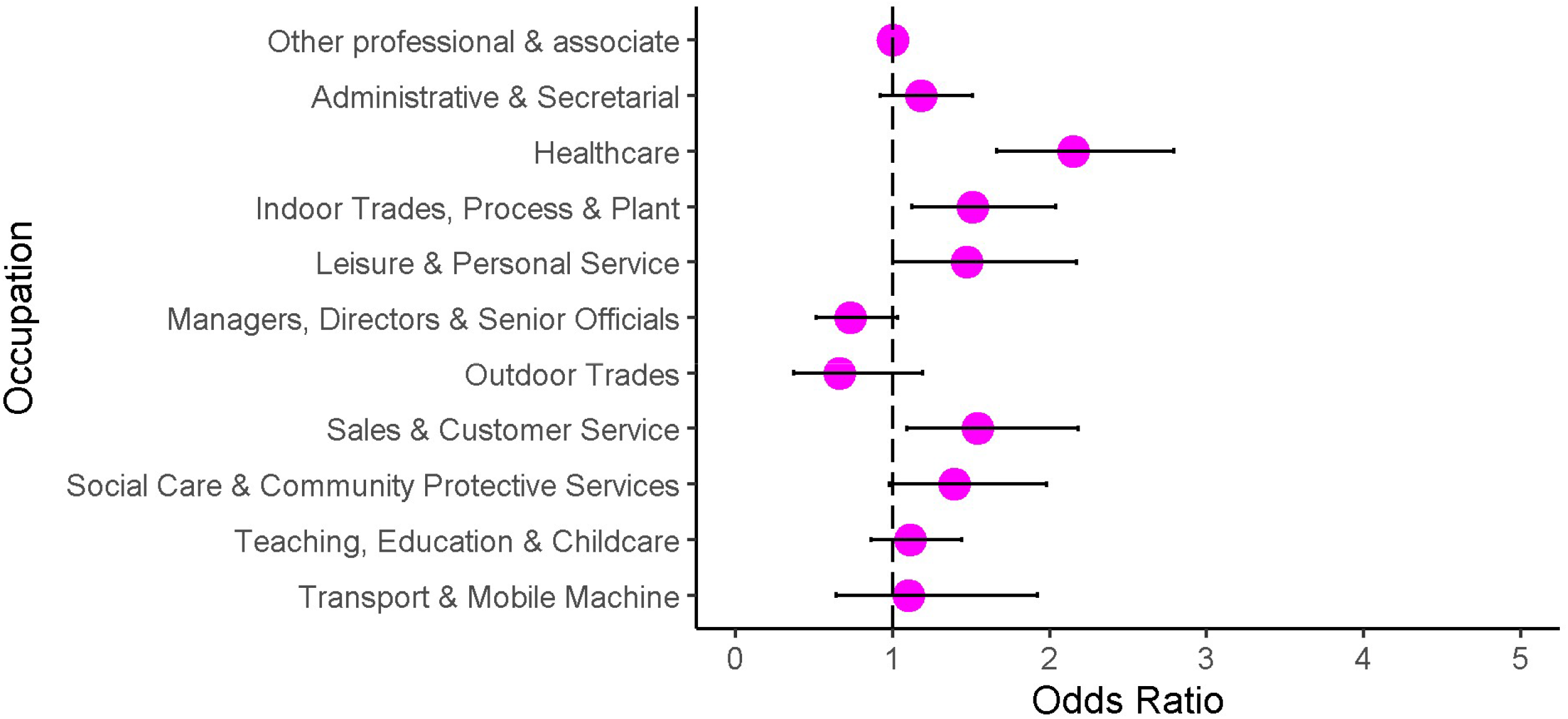
Odds Ratios for Frequency of Exposure to Poorly Ventilated Workplace by Occupation.

## Discussion

Findings from this prospective community cohort study in England and Wales indicated that healthcare workers, indoor trade, process and plant workers, leisure and personal service workers, and transport and mobile machine operatives had at least twice the total odds of seropositivity compared to participants employed in other professional and associate occupations, adjusted for age, sex, household income, and national region. Anti-nucleocapsid seropositivity was highest amongst those with frequent close contact at work (16%) and lowest in those who worked from home or never had frequent close contact at work (10%). Frequency of work-related close contact explained a variable but meaningful part of the increased odds of infection in high-risk occupational groups. After accounting for workplace close contact, healthcare workers and indoor trade, process and plant workers had residual increased odds of infection, suggesting that other work-related factors also contribute to their increased risk^32^. Healthcare workers and indoor trade, process and plant workers also had greater odds of reporting frequent exposure to poor indoor ventilation at work. Mediation models based on observational data must be interpreted with caution in relation to causal inference (see further discussion in Supplementary Materials). Nevertheless, the finding that workplace contact frequency explains a considerable part of the variation in occupational risk is biologically plausible given the transmission routes of SARS-CoV-2^33,34^.

### Strengths and Limitations

Key strengths of the work include objective measurement of prior infection status through anti-nucleocapsid antibody testing which should not be affected by vaccination status. This in effect provides a cumulative measure of the infection risk through the first and second waves of the pandemic in England and Wales. Combining this data on occupational status and potential socio-demographic confounders allowed us to assess the independent effect of occupation on odds of SARS-CoV-2 infection, with adjustment informed by our directed acyclic graph. Furthermore, through use of a mediation model we were able to investigate a putative mechanism for increased occupational risk - work related close contact. Finally, we were able to investigate variations in exposure to poorly-ventilated workplace settings across occupational groups.

Key limitations include that the timing of infection cannot be assessed from the serological tests, and consequently that we cannot determine which infections occurred in the first and second wave. Antibody waning may also lead to false negative results^11^, particularly for infections acquired early in the pandemic. Frequency of work-related contact was also measured during the second pandemic wave and may have changed compared to during the first wave. For many occupational groups - with the notable exception of primary and secondary teaching and education - legislation and guidance around workplace closures was broadly similar across ‘lockdown’ periods with the highest levels of SARS-CoV-2 transmission; consequently, we still considered this item a useful measure of contact frequency for the present study. Work-related contact frequency was self-reported in broad categories and our measure could not account for specific features of contact, such as the number of close contacts across a workday or the duration of contact episodes. Neither could it distinguish between those who worked from home and those who never have close contacts at work. Likewise, we could not assess the presence or impact of risk mitigation methods, such as personal protective equipment (PPE), during close contact. This is likely to mean that we have not fully accounted for risk-relevant features of work-related contact. Nevertheless, our findings suggest that it is an important explanatory variable for differential infection risk across occupational groups. We were not able to directly explore what accounted for the residual risk in healthcare workers and indoor trade and process and plant workers due to lack of statistical power, but nosocomial transmission and poor ventilation during periods of exposure are plausible mechanisms. Relatively small sizes of some occupational groups likely impacted the precision of estimates. Some covariates required broad categorisation to retain statistical power, and household income - while representing a household and not area-level measure of deprivation - may not capture all relevant aspects of deprivation. Reporting of workplace contact frequency and ventilation may have been affected by recall bias. Detailed features and effectiveness of mechanical ventilation are likely to be difficult for a non-specialist to assess.

Further, despite using a directed acyclic graph, to inform sociodemographic confounder adjustment, the complex interrelationships between these factors make excluding these effects challenging. Notably, the relationship between occupation, workplace contact, and serological status may be confounded by occupation-related non-workplace contacts, e.g., using public transport to reach work and contacts outside the workplace that may be increased through attending work. We also did not control for vaccination, although the timing of the antibody tests was such that, other than for healthcare workers, most of those in working age groups will not have been vaccinated. Our mediation model was constrained by statistical power and lack of available data on other relevant workplace factors, such as crowding, ventilation during periods of contact, and PPE.

Finally, it should be noted that this analysis covers periods of time of intense workplace restrictions for many occupations. Occupational patterns are likely to change considerably as restrictions are lifted.

### Interpretation

The elevated total odds of seropositivity identified in the present study among healthcare workers, indoor trade, process and plant occupations, leisure and personal service occupations, and transportation occupations broadly corroborates previous findings for similar occupational groups in the UK and worldwide, indicating elevated risk of infection ^17,19–23^ when compared to the general population or other - usually non-public facing - occupations. These findings support and build on the important role of work-related contact suggested in the ONS Coronavirus Infection survey^24^. Differential infection risk influenced by work-related contact could plausibly contribute to variations in occupational morbidity and mortality observed in other studies ^5–10^, and direct investigation is indicated.

Working from home may eliminate or dramatically reduce this important aspect of occupational risk and is likely to have made a notable contribution to reducing infections during lockdown periods. However, differential ability to work from home will have exacerbated occupational and social inequalities. The extent to which work from home should continue to be encouraged as other restrictions are lifted is an important consideration for society globally. Reducing footfall and maintaining social distancing in the workplace may also be important. The relative importance of these measures will depend on infection levels, vaccination levels and the effectiveness of vaccines against current and future variants of SARS-CoV-2.

High risk in healthcare workers is well-described previously ^11–18^, though accounting for variation between specific occupations and over time due to changing PPE provision and infection control practices were beyond the scope of this study. To our knowledge, the relatively high reported levels of poor ventilation in healthcare settings has not been widely reported within population-based studies, and measures to improve ventilation are likely to be important for control of nosocomial transmission of SARS-CoV-2 and other respiratory infections. Residual risk in indoor trade and process and plant workers, combined with greater reported exposure to poor ventilation, also represents an important area for further investigation and modification to reduce risk. The extent and effectiveness of ventilation is likely to vary considerably according to the design of such workplaces.

## Conclusion

This study is amongst the first to be able to compare occupational risk of SARS-CoV-2 infection after controlling for a range of socio-demographic confounders, and indicates that occupation is an important independent predictor of SARS-CoV-2 infection. Frequency of close contact at work is suggested to explain a considerable amount of this variation. Reducing work-related close contact through measures such as social distancing and working from home is likely to have played an important role in controlling COVID-19 transmission. Poor ventilation in some workplace settings may also contribute to risk, and presents an important area for further inquiry.

## Data Availability

We aim to share aggregate data from this project on our website and via a "Findings so far" section on our website - https://ucl-virus-watch.net/. We will also be sharing individual record level data on a research data sharing service such as the Office of National Statistics Secure Research Service. In sharing the data we will work within the principles set out in the UKRI Guidance on best practice in the management of research data. Access to use of the data whilst research is being conducted will be managed by the Chief Investigators (ACH and RWA) in accordance with the principles set out in the UKRI guidance on best practice in the management of research data. We will put analysis code on publicly available repositories to enable their reuse.

