## Supplementary Materials for "Occupation, Work-Related Contact, and SARS-CoV-2 Anti-Nucleocapsid Serological Status: Findings from the Virus Watch prospective cohort study"

### Discussion of Mediation Models

Total-effect models differ fundamentally from mediation models, and the absence of a total effect does not necessarily indicate the absence of theoretically-sound mediated effects, though these should be interpreted cautiously [1,2]. Following Zhao et al.'s typology of mediation effects [1], our findings suggested indirect-only mediation for all occupational groups except for healthcare workers and indoor trade, process and plant occupations. Direct effects were observed in the mediation model for the latter groups, consistent with complementary mediation[34] and suggesting the presence of further indirect effects unaccounted for in the model.


1 Zhao X, Lynch JG, Chen Q. Reconsidering Baron and Kenny: Myths and Truths about Mediation Analysis. *J Consum Res* 2010; **37**: 197–206.

2 Preacher KJ. Advances in mediation analysis: a survey and synthesis of new developments. *Annu Rev Psychol* 2015; **66**: 825–52.

**Supplementary Figure 1. Directed Acyclic Graph for Estimating the Total and Contact-Mediated Effect of Occupation on SARS-CoV-2 Infection Risk**

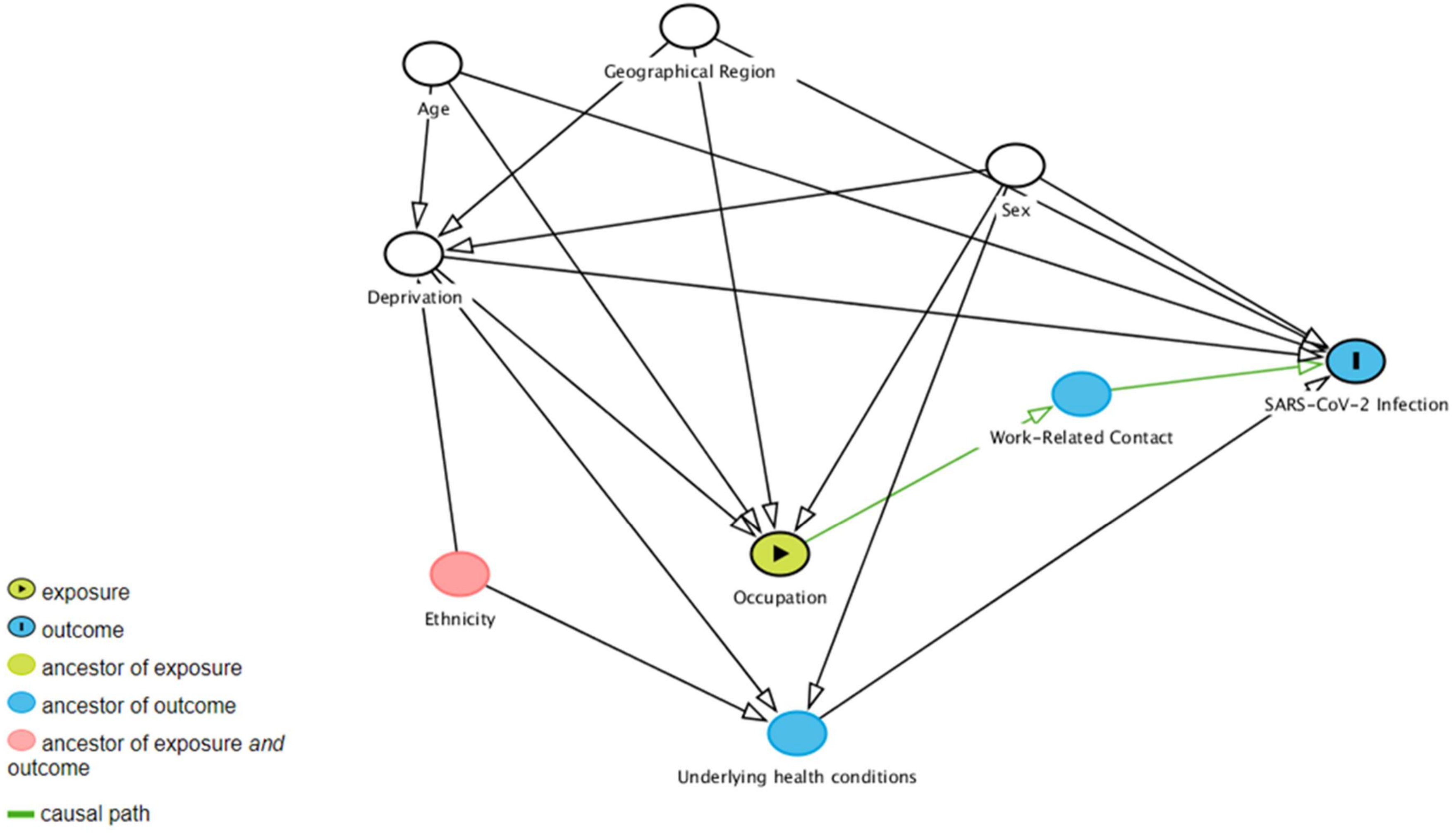

**Supplementary Table 1. UK Standard Occupational Classification 2020 (SOC-2020) Codes within Virus Watch Occupational Categories**

| Virus Watch Occupational Category | UK SOC-2020 Codes | Three Most Prevalent Occupations* (SOC-2020 Unit Group) |
|---|---|---|
| Administrative & Secretarial Occupations | 4111-4217, 9211, 9219, 9233 | 1. Other administrative occupations n.e.c. (24%, n= 118)<br>2. Book-keepers, payroll managers, and wage clerks (11%, n=54)<br>3. Personal assistants and other secretaries (7%, n=35) |
| Healthcare Occupations | 2211-2259, 3211-3219, 6131-6133 | 1. Other nursing professionals (20%, n=65)<br>2. Generalist medical practitioners (10%, n=31)<br>3. Nursing auxiliaries and assistants (7%, n=22) |
| Indoor Trades, Process & Plant Occupations | 5211-5250, 5315-5317, 5321-5323, 5411-5449, 8111-8149, 8160, 9131-9139, 9241-9259 | 1. Metalworking production and maintenance fitters (11%, n=26)<br>2. Warehouse operatives (11%, n=26)<br>3. Electricians and electrical fitters (9%, n=22) |
| Leisure & Personal Service Occupations | 1221-1225, 1252, 1253, 1256, 1257, 6121, 6129, 6211-6250, 9221-9229, 9231, 9262 | 1. Cleaners and domestics (15%, n=22)<br>2. Hairdressers and barbers (10%, n=14)<br>3. Kitchen and catering assistants (7%, n=10) |
| Managers, Directors & Senior Officials | 1111-1161, 1171,1172, 1211, 1212, 1231, 1241-1243, 1255, 1258 | 1. Financial managers and directors (21%, n=57)<br>2. Functional managers and directors n.e.c. (11%, n=31)<br>3. Human resource managers and directors (11%, n=29) |

| | | |
|---|---|---|
| Other Professionals & Associate Professionals | 2111-2162, 2411-2455, 2471-2494, 3111-3133, 3411-3582 | 1. Programmers and software development professionals (6%, n=80)<br>2. Management consultants and business analysts (4%, n=56)<br>3. Business and financial project management professionals (4%, n=51) |
| Outdoor Trade Occupations | 5111-5119, 5311-5314, 5319, 5330, 8151-8159, 9111- 9129 | 1. Gardeners and landscape gardeners (27%, n=24)<br>2. Farmers (18%, n=16)<br>3. Construction operatives n.e.c. (13%, n=12) |
| Sales & Customer Service Occupations | 7111-7220 | 1. Sales and retail assistants (36%, n=63)<br>2. Customer service occupations n.e.c. (13%, n=22)<br>3. Sales supervisors - retail and wholesale (11%, n=20) |
| Social Care & Community Protective Services | 1162, 1163, 2461-2669, 3221-3229, 3311-3319, 6134-6138, 6311-6312 | 1. Care workers and home carers (26%, n=47)<br>2. Welfare and housing associate professionals n.e.c. (12%, n=22)<br>3. Social workers (9%, n=16) |
| Teaching, Education & Childcare Occupations | 2311-2329, 3231, 3232, 6111-6117, 9232 | 1. Higher education teaching professionals (14%, n=64)<br>2. Education advisers and school inspectors (13%, n=58)<br>3. Secondary education teaching professionals (13%, n=58) |
| Transport & Mobile Machine Operatives | 8211-8239 | 1. Large goods vehicle drivers (23%, n=18)<br>2. Bus and coach drivers (14%, n=11)<br>3. Driving instructors (14%, n=11) |

**Abbreviations:** n.e.c. = not elsewhere classified

* Limited to three most prevalent occupations per category to prevent declarative disclosure and due to large number of occupations across sample (*n*=363

**Supplementary Table 2. Frequency of Workplace Close Contact and Seropositivity by Occupation**

| Characteristic | | Daily, N = 707[1] | Intermediate, N = 933[1] | Never, N = 2,121[1] |
|---|---|---|---|---|
| Occupation | Other Professional & Associate | 88.0 (6.7%) | 238.0 (18.1%) | 987.0 (75.2%) |
| | Administrative & Secretarial | 68.0 (13.7%) | 125.0 (25.2%) | 303.0 (61.1%) |
| | Healthcare | 135.0 (42.3%) | 108.0 (33.9%) | 76.0 (23.8%) |
| | Indoor Trades, Process & Plant | 79.0 (32.0%) | 84.0 (34.0%) | 84.0 (34.0%) |
| | Leisure & Personal Service | 40.0 (27.8%) | 43.0 (29.9%) | 61.0 (42.4%) |
| | Managers, Directors & Senior Officials | 39.0 (14.3%) | 52.0 (19.0%) | 182.0 (66.7%) |
| | Outdoor Trades | 24.0 (26.7%) | 33.0 (36.7%) | 33.0 (36.7%) |
| | Sales & Customer Service | 39.0 (22.3%) | 53.0 (30.3%) | 83.0 (47.4%) |
| | Social Care & Community Protective Services | 43.0 (23.9%) | 61.0 (33.9%) | 76.0 (42.2%) |
| | Teaching, Education & Childcare | 119.0 (26.7%) | 111.0 (24.9%) | 216.0 (48.4%) |
| | Transport & Mobile Machine | 33.0 (42.3%) | 25.0 (32.1%) | 20.0 (25.6%) |
| Anti-Nucleocapsid Seropositive | Yes | 113 (16.0%) | 120 (12.9%) | 203 (9.6%) |
| | No | 594 (84.0%) | 813 (87.1%) | 1918 (90.4%) |

[1]n (%)

**Supplementary Table 3. Frequency of Exposure to Poorly Ventilated Workplace by Occupation**

| Characteristic | | **Daily, N = 401**[1] | **Intermediate, N = 497**[1] | **Never, N = 2,836**[1] |
|---|---|---|---|---|
| | Other Professional & Associate | 133 (10.2%) | 140 (10.7%) | 1,034 (79.1%) |
| | Administrative & Secretarial | 56 (11.4%) | 61 (12.4%) | 375 (76.2%) |
| | Healthcare | 50 (15.9%) | 68 (21.6%) | 197 (62.5%) |
| | Indoor Trades, Process & Plant | 28 (11.4%) | 44 (18.0%) | 173 (70.6%) |
| | Leisure & Personal Service | 22 (15.5%) | 17 (12.0%) | 103 (72.5%) |
| | Managers, Directors & Senior Officials | 20 (7.4%) | 24 (8.9%) | 227 (83.8%) |
| | Outdoor Trades | 3 (3.3%) | 11 (12.2%) | 76 (84.4%) |
| | Sales & Customer Service | 23 (13.2%) | 28 (16.1%) | 123 (70.7%) |
| | Social Care & Community Protective Services | 20 (11.2%) | 29 (16.3%) | 129 (72.5%) |
| | Teaching, Education & Childcare | 37 (8.3%) | 67 (15.1%) | 340 (76.6%) |
| | Transport & Mobile Machine | 9 (11.8%) | 8 (10.5%) | 59 (77.6%) |
| Anti-Nucleocapsid Seropositive | Yes | 67 (16.7%) | 62 (12.5%) | 303 (10.7%) |
| | No | 334 (83.3%) | 435 (87.5%) | 2533 (89.3%) |

[1] n (%)

**Supplementary Table 4. Odds Ratios for Frequency of Exposure to Poorly Ventilated Workplace by Occupation**

| | OR | 95% CI | *p* |
|---|---|---|---|
| Other Professional & Associate | REF | REF | REF |
| Administrative & Secretarial | 1.18 | 0.92,1.51 | 0.19 |
| Healthcare | 2.15 | 1.66,2.79 | <0.001 |
| Indoor Trades, Process & Plant | 1.51 | 1.12,2.04 | 0.01 |
| Leisure & Personal Service | 1.47 | 1.00,2.17 | 0.05 |
| Managers, Directors & Senior Officials | 0.73 | 0.51,1.03 | 0.07 |
| Outdoor Trades | 0.66 | 0.37,1.19 | 0.17 |
| Sales & Customer Service | 1.54 | 1.09,2.18 | 0.01 |
| Social Care & Community Protective Services | 1.39 | 0.98,1.98 | 0.06 |
| Teaching, Education & Childcare | 1.11 | 0.86,1.44 | 0.41 |
| Transport & Mobile Machine | 1.10 | 0.64,1.92 | 0.72 |
